# Exploring the Psychological, Social, and Human Rights Dynamics and Consequences of COVID-19 Control Measures: A Systematic Review

**DOI:** 10.1101/2024.03.18.24304509

**Authors:** Djurdje Spasojevic

## Abstract

This systematic review explores the social and psychological dynamics and consequences of COVID-19 control measures and their implications for human rights. Through a lens of social psychology, the review considers factors such as social influence, obedience, perceived control, social comparison, cognitive dissonance, propaganda, surveillance, fearmongering, incentives, coercion, persuasion, censorship, obfuscation, isolation, and rewards and punishment. By analyzing the influence of these factors on individuals and group responses to the pandemic and the manipulation of social and psychological dynamics by institutions to shape compliance, this review provides insights into the determinants that drive adherence to control measures and their negative consequences. The findings of the 13 selected studies contribute to understanding the multifaceted factors that influence compliance and inform the development of effective public health interventions to avoid consequences. The review emphasizes the importance of upholding human rights during the implementation of control measures, given the reported violations across the world. By providing insights to policymakers, politicians, health practitioners, and researchers, this review enables the formulation of strategies that promote public health while respecting human and individual rights and well-being. In conclusion, this study sheds light on the social and psychological dynamics, human rights, and implications of COVID-19 control measures, providing valuable insights for future interventions.

## Introduction

The COVID-19 pandemic has had far-reaching social and psychological consequences, primarily attributed to the implementation and promotion of control measures by various entities, such as governments, media, and international institutions. This systematic review aims to comprehensively examine the social and psychological effects of COVID-19 control measures, particularly focusing on potential human rights implications. The theoretical framework guiding this review is rooted in social and group psychology, drawing upon key concepts, including group psychology, social learning theory, cognitive dissonance, groupthink (crowd psychology), obedience to authority, and learned helplessness.

### Theoretical Framework

The theoretical framework for understanding societal responses to COVID-19 control measures and their prospective misuse can be organized into three categories: the influence of group dynamics, the impact of coercive tactics and obedience to authority, and individual psychological responses.

#### Group Dynamics

Group psychology and group dynamics have emerged as crucial concepts that help understand individual responses to pandemic control measures and their misuse. According to Social Identity Theory (1), individuals’ behavior during the COVID-19 pandemic is significantly shaped by their affiliation and identification with social groups. Individuals form an identity based on their group memberships and shape their behavior, attitudes, and beliefs. This group identity significantly impacts adherence to control measures such as mask-wearing, physical distancing, and quarantining, even if detrimental to individuals’ psychological and social well-being and rights.

Bandura’s (2) Social Learning Theory posits that people acquire behaviors through observation, imitation, and vicarious reinforcement. Applied to the pandemic context, this theory helps explain how individuals learn to comply with control measures by observing others’ behavior or fostering feelings of shame, persuasion, or coercion. Literature indicates that social norms and social pressure play a significant role in shaping compliance measures (3).

Furthermore, the dynamics of groupthink (4), also known as crown psychology, is a psychological phenomenon that significantly affects behavior. Group members tend to belong and often neglect their individual feelings in the process and prioritize consensus and conformity over individual critical thinking and decision making. It is essential to examine the groupthink dynamics in the development and implementation of pandemic control measures. Understanding the influence of group dynamics on decision-making processes and groupthink’s potential to shape pandemic responses provides valuable insights into social and psychological factors at play.

#### Coercive Tactics and Obedience to Authority

Understanding societal responses also involves analyzing the persuasive and coercive tactics employed during the pandemic and the inherent obedience to authority. Tactics such as propaganda, surveillance, fearmongering, nudges, incentives, censorship, and obfuscation have been used to manipulate and coerce behavior and compliance with control measures (5). These tactics aim to shape public perceptions, attitudes, and compliance with the prescribed control measures.

One landmark study that sheds light on obedience to authority is Milgram’s (6) study. Milgram investigated how ordinary individuals could be influenced to commit morally questionable acts in the direction of an authority figure. Participants were asked to administer electric shocks to another individual (actor) when they answered questions incorrectly. Despite the actor’s apparent discomfort and pleas for the experiment to stop, a significant portion of the participants obeyed the authority figure’s instructions and continued to administer increasingly severe shocks. This study demonstrates the profound impact of authority on individuals’ behavior, highlighting the potential for coercion and obedience in the context of COVID-19 control measures and their implications for human rights.

#### Psychological Responses

Cognitive Dissonance Theory (7) captures the mental discomfort experienced by individuals when they hold conflicting beliefs or engage in behavior that contradicts these beliefs. This theory helps conceptualize how individuals reconcile conflicting beliefs and behaviors. For instance, individuals valuing their freedom may experience cognitive dissonance when they are required to adhere to measures such as mask-wearing or lockdown orders that seemingly restrict their autonomy. The literature on Social Comparison Theory (8) provides a lens for examining how individuals compare themselves to others in terms of pandemic control measures. This theory suggests that individuals evaluate their abilities, opinions, and social standing by comparing themselves to others.

Finally, the concept of Learned Helplessness (9) sheds light on the psychological and social effects of COVID-19 control measures. This phenomenon occurs when people perceive that they have no control over their environment, which leads to passivity and apathy in the face of adverse events. The prolonged nature of the pandemic, the uncertainty surrounding its resolution, and the impact of control measures on mental health, particularly for individuals experiencing multiple lockdowns, prolonged periods of isolation, and restrictive measures, may lead to a sense of helplessness and further psychological distress. Learned helplessness, as demonstrated in the study by Abramson et al.(10), has been linked to negative outcomes such as depression, anxiety, and reduced motivation.

This theoretical framework seeks to provide a comprehensive understanding of the complex interplay of psychological responses, such as group dynamics, coercive tactics, obedience to authority, and individual psychological responses during the COVID-19 pandemic and the broader societal implications during and after the pandemic.

### Studies and Reports

In addition to psychological and social consequences, COVID-19 control measures can have significant social and economic impacts, which further deteriorate the well-being of individuals and groups. Certain groups, such as low-income individuals who could not work from home and had to continue to expose themselves to risks, have been disproportionately affected by lockdowns and other control measures. The economic consequences of the pandemic and its associated control measures have exacerbated existing inequalities, amplifying their adverse effects on vulnerable populations.

The article titled "Covid-19: Politicisation, "Corruption," and Suppression of Science" by Abbasi K. (11) in the British Medical Journal highlights how political pressures and conflicts of interest have resulted in the suppression and distortion of science during the COVID-19 pandemic. The author argues that these actions have led to avoidable deaths, increased economic and social disruption, and undermined public confidence in government and science. Abbassi called for an end to the pandemic politics and urged that public health policy should be guided by science and transparency.

In the study conducted by Singh et al. (12), the authors investigated the negative consequences of school closures during the COVID-19 pandemic on children and adolescents in India. The findings suggest that extended school closures not only disrupted education but also led to increased rates of child labor, malnutrition due to missing school meals, and exacerbations of social inequities. The authors called for strategies to mitigate these effects, underscoring the need for child-focused interventions during pandemics.

An article by Gostin et al. (13) published in The Lancet provides an analysis of human rights violations during the COVID-19 pandemic. The authors emphasize that many states have failed to fulfill their human rights obligations throughout the pandemic. They highlighted the initial suppression of information by Chinese officials in Wuhan, which violated freedom of expression and the right to health. The effects of the pandemic have also revealed profound inequalities, both within countries and globally, exposing the failure to achieve non-discrimination and the highest attainable standard of health for all. This study proposes embedding human rights and equity within a transformed global health architecture as a necessary response to address these violations. The authors call for the integration of human rights into a new pandemic treaty, and the establishment of new legal instruments and mechanisms to promote equality and human rights in future health emergencies.

Amnesty International reported police abuse in enforcing COVID-19 restrictions, including excessive force and targeting minorities in countries like Angola, El Salvador, Kenya, etc. The report also addresses prison riots in nations such as Thailand and the USA, discriminatory practices against Roma in Bulgaria and Slovakia, and restrictions on asylum seekers in Cyprus. Additionally, it highlights the misuse of misinformation laws to silence critics in Turkey and Hungary, raising concerns over the erosion of freedoms under the guise of pandemic control (14).

Kelly Delvac highlights global human rights abuses under COVID-19 containment efforts, underscoring the balance between public health and individual freedoms. She points out the punitive enforcement measures in countries like the Philippines, Brazil, and South Africa, which she argues contravene the U.N. Universal Declaration of Human Rights. Delvac advocates for U.N. actions against these violations, spotlighting the pandemic’s exposure of entrenched issues of police brutality and systemic inequality (15).

The WHO highlights a 25% rise in global anxiety and depression due to COVID-19, stressing the impact on youth, women, and those with health conditions. The pandemic has intensified mental health challenges, revealing significant care gaps and the critical need for increased mental health investment and support. Urging international action, the WHO calls for a global enhancement of mental health services to address the pandemic’s widespread psychosocial effects (16). Similarly, in the United Kingdom, critiques have been leveled against the government for its management of the pandemic and the impact of containment measures on mental health, social well-being, and human rights (17).

UN Secretary-General António Guterres warned that the COVID-19 pandemic has been used as a pretext in many countries to crush dissent, criminalize freedoms, and silence reporting, marking a global surge in human rights abuses. This emphasizes a critical global issue, aligning with concerns in various nations over the application of emergency powers to enforce control, often through excessive measures (18). Human Rights Watch has documented numerous cases of authorities exploiting the pandemic to justify excessive force, arbitrary detention, and other rights violations (19). The WHO has expressed concerns about human rights violations associated with COVID-19 control measures in various countries, including Cambodia, Ethiopia, and Uganda, where excessive force and arbitrary detention have apparently been used to enforce lockdowns and curfews (20).

The call from the UN High Commissioner for Human Rights to ensure the implementation of pandemic control measures in a manner that upholds human rights and the need for accountability in cases of abuse or violations underscores the importance of protecting individual rights during a COVID-19 pandemic (21). These examples highlight the potential for political and institutional abuse and exploitation, emphasizing the necessity for ongoing monitoring and corrective action to safeguard human rights.

The COVID-19 pandemic has served as a reminder of the significance of human rights and the potential violations that may occur during the implementation of control measures. The United Nations stressed the importance of aligning pandemic control measures with human rights principles such as non-discrimination, equality, and the right to health (22). However, it is essential to acknowledge that some measures may encroach on individual rights and freedoms. Documented instances of rights violations underscore the need for a nuanced approach to balancing public health imperatives with individual liberties.

Balancing public health objectives with the protection of individual rights and freedoms is a complex task, necessitating careful consideration and respect for human rights principles. This review utilizes a theoretical framework rooted in social psychology to examine how social norms, group dynamics, and individual factors influence compliance with pandemic control measures globally, as well as their misuse and exploitation by governments and other institutions. It also assesses the broader societal and global impacts of these measures, including their contribution to and deterioration of social and psychological realities. The findings offer valuable insights into the effects of COVID-19 control measures, informing future policymaking processes.

### Rationale

The COVID-19 pandemic has posed unprecedented challenges to global public health with significant social, psychological, and economic consequences. The implementation of control measures to manage crises has raised concerns about their impact on human rights and the potential for violations. A systematic review is necessary to address these concerns and gain a comprehensive understanding of the social and psychological dynamics surrounding COVID-19 control measures.

Furthermore, it is crucial to examine the implementation of control measures, their consequences, and their capacity for institutional abuse and exploitation. Reports of excessive force, arbitrary detention, and other human rights violations emerged in various countries during the pandemic. Understanding these and their social and psychological ramifications is essential to ensure justice and future control measures should be implemented in a manner that respects human rights and minimizes adverse consequences.

Through a systematic review that synthesizes, critically analyzes, and evaluates existing peer-reviewed studies, this study, even though limited in scope because of the tremendous complexity of the issue at hand, aims to provide a comprehensive understanding of the social and psychological factors that influence compliance with COVID-19 control measures and its human rights implications.

### Aims and Objectives

This systematic review aims to examine the influence of psychological, social, and group dynamics on compliance with pandemic control measures and to investigate the human rights implications of coercive tactics used during the pandemic. To achieve this aim, the following objectives were addressed: (1) to critically evaluate the existing literature on social influence, obedience to authority, perceived control, social comparison, cognitive dissonance, rewards and punishment, isolation, etc., and their effects on compliance with pandemic control measures; (2) to investigate the implications on human rights of propaganda, surveillance, fear-mongering, incentives, censorship, obfuscation, etc., and their use during the pandemic; (3) to examine how group dynamics and social identity shape people’s responses and behavior to the pandemic, including how social norms and social pressure influence compliance with pandemic control measures; and (4) to identify any gaps in the literature and highlight areas for future research. The findings of this systematic review will contribute to a better understanding of human rights abuses, social and psychological consequences, and factors that influence compliance with pandemic control measures and inform future public health interventions.

### Research Questions

1. How do social and psychological factors influence adherence to COVID-19 control measures?
2. What are the reported cases of human rights violations associated with the implementation of COVID-19 control measures in different countries?
3. How do social norms, social pressure, and tactics, such as propaganda, surveillance, fearmongering, incentives, censorship, and obfuscation, influence compliance with the COVID-19 control measures?
4. What gaps exist in the current literature on the social, psychological, and human rights impacts of the COVID-19 control measures?

## Methods

In line with the SPIDER framework (Table 1), this review acknowledges the potential of encountering studies that possess overlapping, but not entirely identical, focus areas. Determining how to handle these studies requires careful consideration. Defined inclusion and exclusion criteria, which delineate the scope of the review and identify relevant studies, were established from the outset. For studies that may not align exactly with the core research questions but hold an overlapping focus, an evaluation will be undertaken to assess whether they can provide valuable insights into the wider understanding of the social and psychological dynamics of COVID-19 control measures and their human rights implications.

**Table 1.** SPIDER framework.

| SPIDER Framework | Description |
| --- | --- |
| S - Sample | Sample characteristics: The target population for this systematic review includes groups affected by COVID-19 control measures, such as mask-wearing, physical distancing, quarantining, lockdowns, vaccine mandates, and other controlled preventive actions. The sample may comprise individuals from diverse countries, socio-economic backgrounds, and demographic groups. |
| PI - Phenomenon of Interest | The phenomenon of interest is the social and psychological dynamics of COVID-19 control measures and their potential impact on human rights. The focus is on understanding factors such as social influence, attitudes, obedience to authority, perceived control, social comparison, cognitive dissonance, propaganda, surveillance, social engineering, fear-mongering, incentives, censorship, confusion, rewards and punishment, and isolation. |
| D - Design | The design of this systematic review involves the collection and analysis of existing literature on the social and psychological dynamics of COVID-19 control measures. The review will include qualitative, quantitative, and mixed-methods studies, as well as reports, articles, and official documents that provide insights into the phenomenon of interest. |
| E - Evaluation Outcome measure | The evaluation outcome measures in this systematic review may include attitudes, beliefs, behaviors, compliance rates, mental health outcomes, reports of human rights violations, and the impact of social and psychological factors on adherence to COVID-19 control measures. |
| R - Research Type | This research can be classified as a mixed-methods systematic review, as it aims to incorporate qualitative and quantitative studies, as well as reports and documents, to comprehensively examine the social and psychological dynamics of COVID-19 control measures and their potential impact on human rights. |

*Note:* The table includes the different components of the SPIDER framework (Sample, Phenomenon of Interest, Design, Evaluation Outcome measure, Research Type), along with their respective descriptions.

The SPIDER framework was chosen for its suitability for addressing complex research questions within systematic reviews of qualitative and mixed-methods research. This framework allows for a nuanced exploration of the sample, phenomenon of interest, design, evaluation, and research type, making it particularly suitable for investigating the multifaceted impact of COVID-19 control measures. Its application facilitated a structured and comprehensive approach to capture and synthesize evidence across diverse study designs, contributing to a deeper understanding of the pandemic’s psychosocial effects and human rights implications.

The quality assessment of each study is integral and will be performed using a dedicated evaluation table. During data extraction and synthesis, care was taken to ensure that only the relevant data were extracted from these studies. The report outlines the approach taken in handling overlapping but not identical studies, supporting an understanding of the decision-making process and paving the way for future replication of the study. This review also discusses how the inclusion of these studies may have affected the findings in the discussion.

In selecting the electronic databases for this systematic review (Table 2), ProQuest, Science Direct (ELSEVIER), PsycINFO (Medline), and PubMed were chosen because of their comprehensive coverage of the health-related and psychological literature. This selection ensured a broad and relevant range of peer-reviewed articles, encompassing the multifaceted implications of COVID-19 control measures on social dynamics, psychological well-being, and human rights. The inclusion and exclusion criteria were meticulously designed to capture the essence of COVID-19’s impact across diverse settings and populations, ensuring a focused yet comprehensive exploration of the pandemic’s social and psychological ramifications. This strategic approach aims to provide a robust synthesis of current knowledge, contributing to the development of informed public health interventions and policymaking.

**Table 2.** Database Searches.

| Database | Search Terms | Filters | Results |
| --- | --- | --- | --- |
| **ProQuest** | (COVID-19 OR coronavirus OR pandemic) AND (Social consequences OR social behavior OR social impact OR social changes) AND (Psychological consequences OR psychological distress OR mental health OR psychological impact) AND (Human rights OR Human rights violations OR Human rights abuses) | Scholarly Journals, 2020-03-01 to 2023-06-09 | 2073 |
| **Science Direct (Elsevier)** | COVID-19 OR coronavirus AND Social consequences OR social impact OR social changes AND Psychological consequences OR psychological distress OR mental health AND Human rights violations | Subscribed journals, Years: 2020-2023, Article type: Review articles, Subject areas: Social Sciences, Psychology, Access type: Open Access & Open archive | 926 |
| **PsycINFO/ PsycNET** | Any Field: "COVID-19" OR Any Field: "coronavirus" OR Any Field: "pandemic" AllFields AND Any Field: Social consequences OR Any Field: social behavior OR Any Field: social impact OR Any Field: social changes AllFields AND Any Field: Psychological consequences OR Any Field: psychological distress OR Any Field: mental health OR Any Field: psychological impact AllFields AND Any Field: Human rights OR Any Field: Human rights violations OR Any Field: Human rights abuses AllFields AND Any Field: "Peer Reviewed Journal" AND Population Group: Human AND Age Group: Adulthood (18 yrs & | APA PsycInfo, APA PsycArticles, APA PsycBooks, APA PsycExtra | 832 |
|  | older) OR Young Adulthood (18-29 yrs) OR Thirties (30-39 yrs) OR Middle Age (40-64 yrs) AND Publication Type: Peer Reviewed Journal AND Index Term: COVID-19 AND Population Group: Human AND Year: 2020 To 2023 |  |  |
| **PubMed - MEDLINE** | ((COVID-19[Title/Abstract] OR coronavirus[Title/Abstract] OR pandemic[Title/Abstract]) AND (Social consequences OR social behavior OR social impact OR social changes)) AND (Psychological consequences OR psychological distress OR mental health OR psychological impact)) AND (Human rights OR Human rights violations OR Human rights abuses) | Humans, MEDLINE, English, from 2020-2023 | 102 |

*Note: Search terms targeted the wide-ranging effects of COVID-19 control measures on societal, psychological, and human rights issues. Filters applied to include pertinent, quality studies from the specific period, focusing on current scholarly discourse.*

### Eligibility Criteria

#### Inclusion Criteria

##### Population

Individuals of all ages and genders who have been directly impacted by COVID-19 control measures (such as quarantine, social distancing, and mask mandates) and specific human rights violations related to these measures.

##### Context

Studies conducted in various settings (e.g., communities, institutions, and countries) where COVID-19 control measures and human rights violations have occurred.

##### Study Design

Primary qualitative studies, including but not limited to ethnographic research, phenomenological studies, and qualitative data within mixed-methods studies. Quantitative studies with specific features (e.g., including what types of quantitative studies should be included).

##### Language

Studies published in English.

##### Research Question

Studies that aim to explore the social and psychological impacts, experiences, perceptions, or responses of individuals or communities concerning COVID-19 control measures and human rights violations.

#### Exclusion Criteria

##### Population

Studies focusing exclusively on specific subpopulations (e.g., healthcare workers, children, specific ethnic groups, individuals with pre-existing mental health conditions) unless they provide insights relevant to a broader understanding of the social and psychological dynamics of COVID-19 control measures and human rights violations.

##### Study Design

Quantitative studies that do not incorporate qualitative data or provide an in-depth exploration of social and psychological experiences.

##### Irrelevant Outcomes

Studies that do not specifically address the social and psychological impacts of COVID-19 control measures and human rights violations (studies focusing solely on economic or political implications).

##### Language

Studies published in languages other than English.

### Information sources

Electronic databases— ProQuest, Science Direct (ELSEVIER), PsycINFO/PsycNET, and PubMed —are chosen for their expansive health-related and psychological literature coverage. Keyword and Boolean operator consistency will be ensured throughout the search strategy. The choice of these databases is based on their alignment with the study’s focus, and their vast array of peer-reviewed articles that offer insights into the human rights and psychosocial implications of COVID-19 control measures.

#### Keywords used

(General Terms, MeSH): COVID-19, pandemic, social dynamics (consequences), psychological dynamics (consequences), control measures, and human rights.

#### General keywords with Boolean operators

("COVID-19" OR "Coronavirus") AND ("social dynamics" OR “social consequences”) AND ("psychological dynamics" OR “psychological consequences”) AND ("control measures" OR "public health measures") AND ("human rights" OR "civil liberties")

### Search strategy

In formulating the search strategy, keyword selection is intentionally comprehensive to account for the multidimensionality of the research question. Selected keywords like "COVID-19", "coronavirus", “pandemic”, "human rights", "freedoms", "control measures", "restrictions", "psychological effects", and "social impact", capture the intersection between the pandemic and human rights discourse and well-being. Utilizing Boolean operators AND/OR allows the search to be broad yet focused, ensuring all pertinent literature is considered while minimizing the inclusion of unrelated material as presented in Table 2.

### Screening of Studies and PRISMA Flowchart

In this study, a systematic and rigorous approach was taken to select relevant studies for inclusion. The screening process was conducted using Covidence, a systematic review management tool, which facilitated efficient and organized screening of studies. Initially, a total of 3931 references were imported for screening. After removing 37 duplicates, 3894 studies were assessed against their titles and abstracts. During this stage, 3563 studies were excluded based on their relevance to the research aims and hypotheses. Subsequently, 331 studies were further assessed for full-text eligibility. Following a thorough evaluation, 318 studies were excluded due to various reasons, such as being irrelevant to the study’s indication, intervention, outcomes, comparator, setting, patient population, or route of administration. Ultimately, 13 studies met the inclusion criteria and were included in the final analysis. This systematic screening process using Covidence ensured a comprehensive and unbiased selection of studies, enhancing the validity and reliability of the findings, which can be seen in Fig 1. The synthesis of results will be presented in the narrative synthesis format.

**Figure 1.**
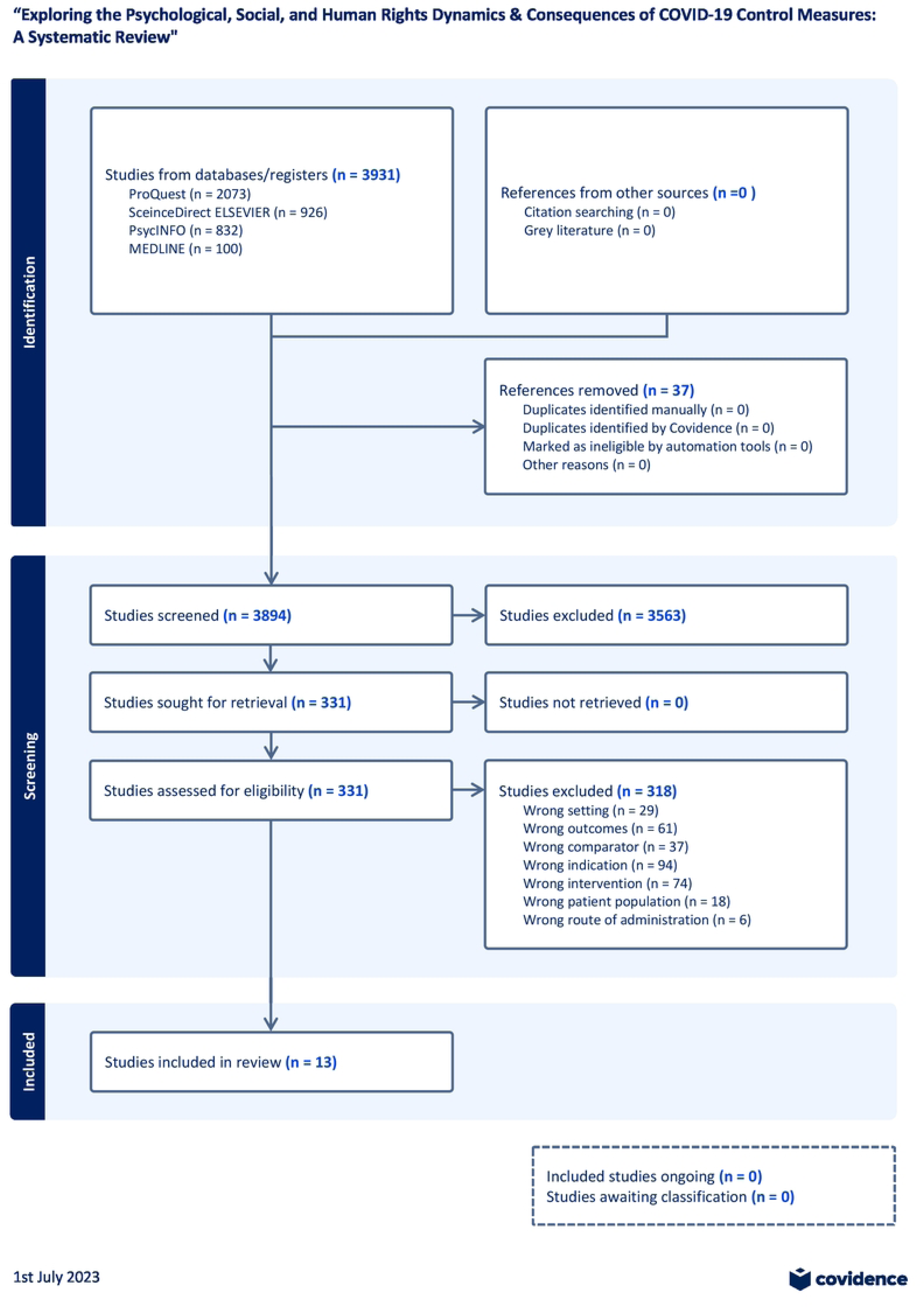

### Risk of Bias (RoB) and Quality Assessment

The assessment of the risk of bias and quality of studies (Table 3) in this systematic review incorporated modified versions of the Cochrane Risk of Bias tool for randomized trials (RoB 2) and the ROBINS-I tool for non-randomized studies (23). These tools were adapted to suit the diverse study types encompassed within the review. The variance in risk of bias and quality of evidence was notable among the 13 studies, driven by their design and type. For instance, quantitative cross-sectional surveys generally displayed low risk of bias and high evidence quality due to their robust methodologies. Commentary articles, which are characteristically subjective, demonstrated a low risk of bias but lower evidence quality. Review articles exhibited varied risks of bias and evidence quality, influenced by potential subjectivity in study selection.

**Table 3.**
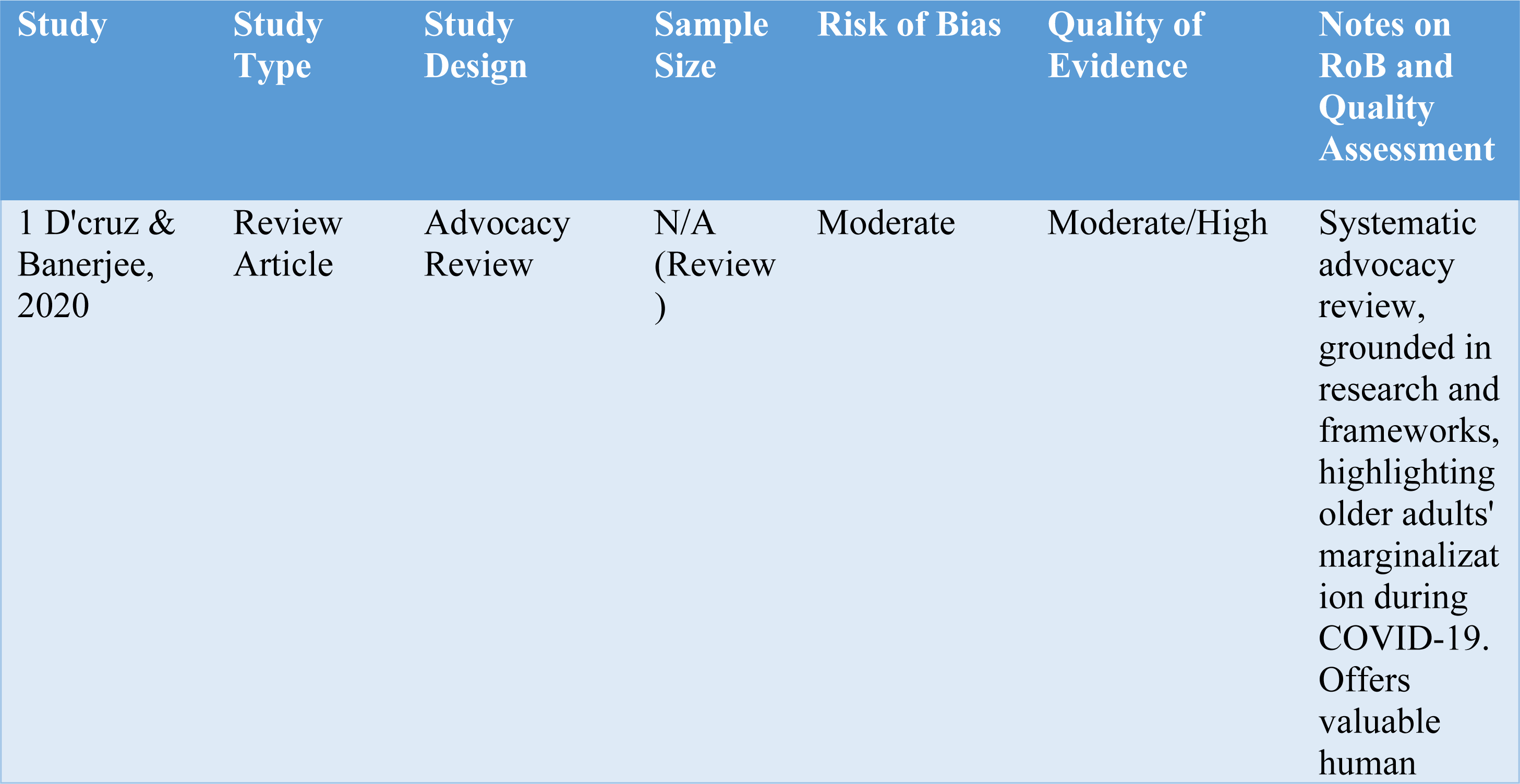
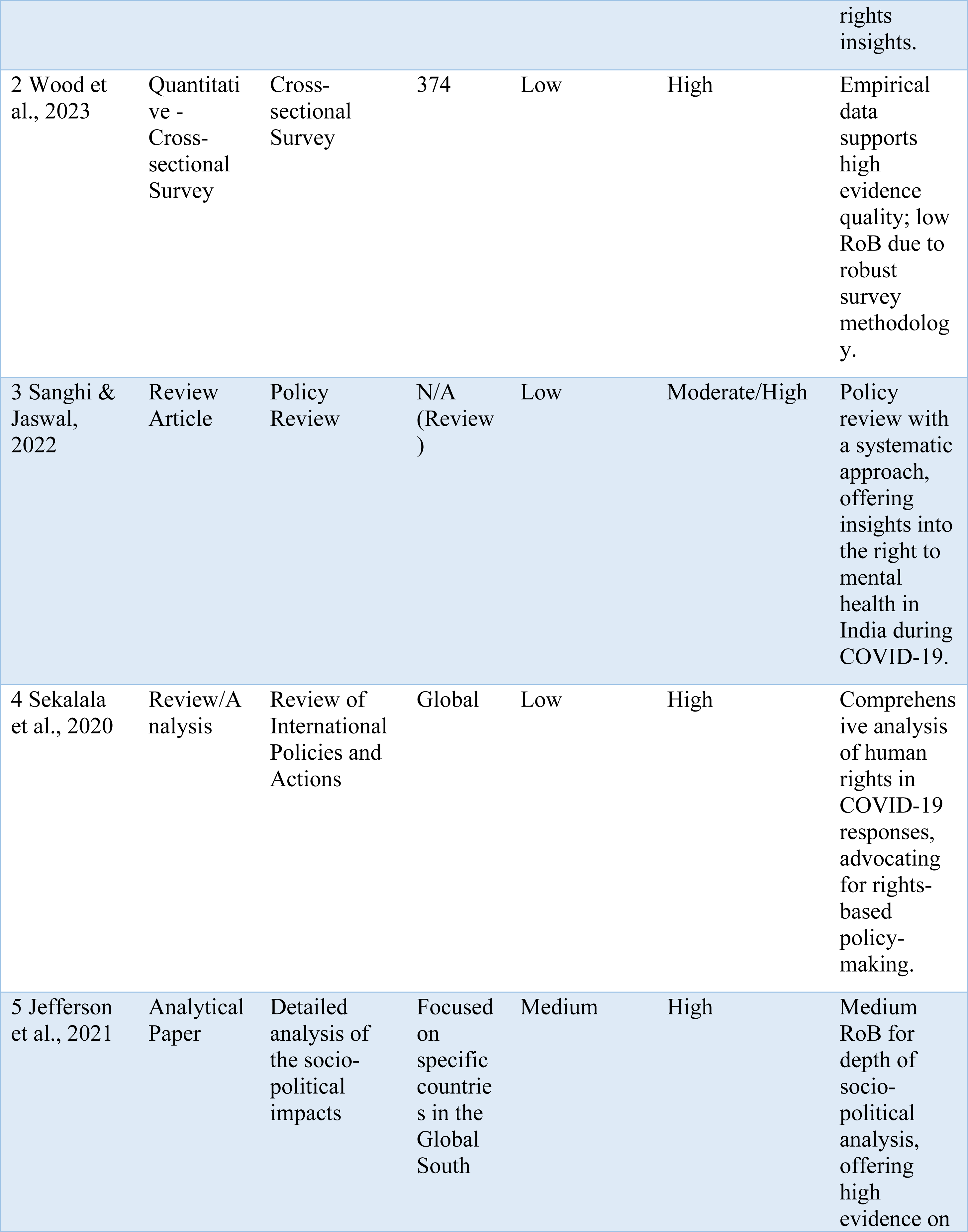
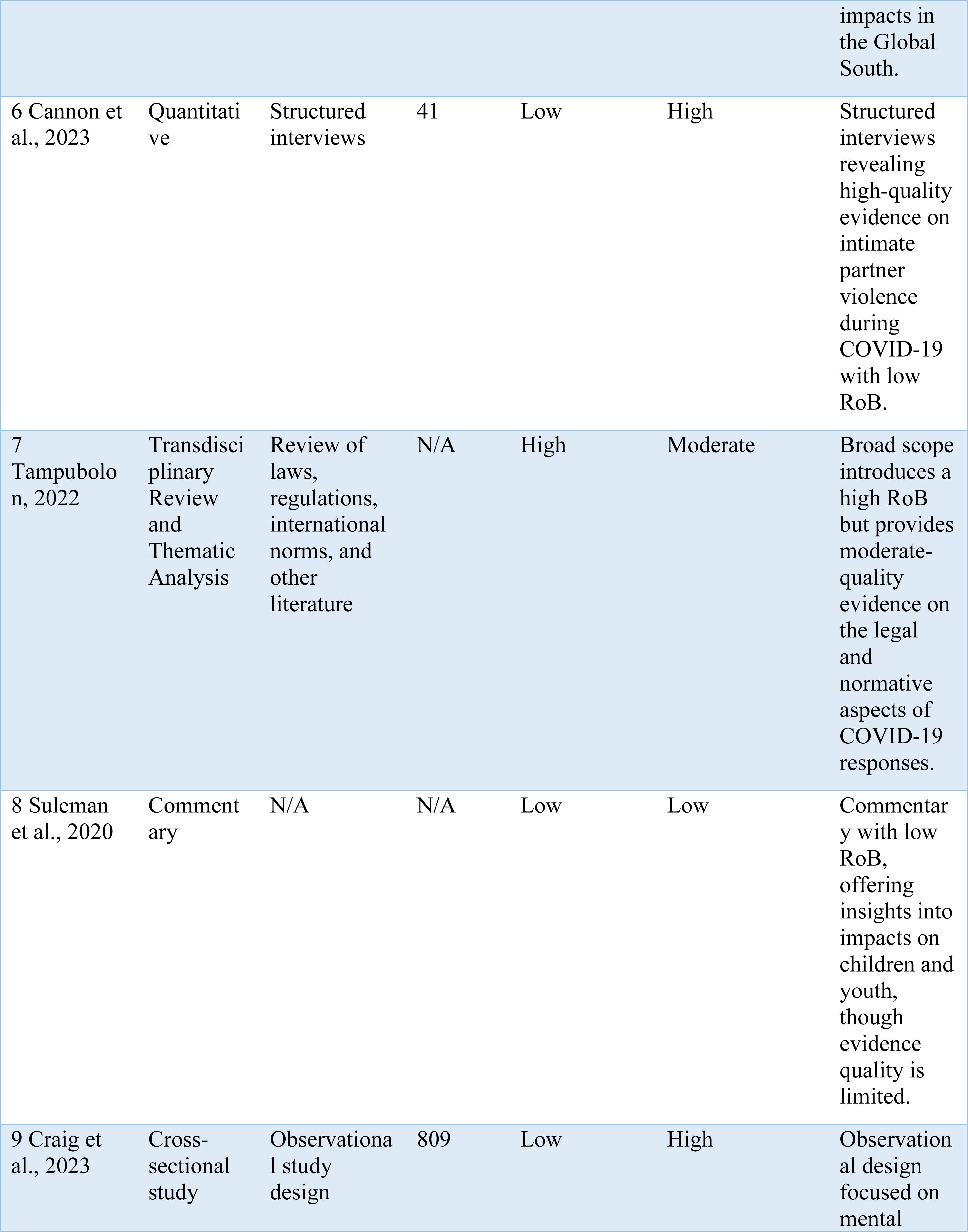
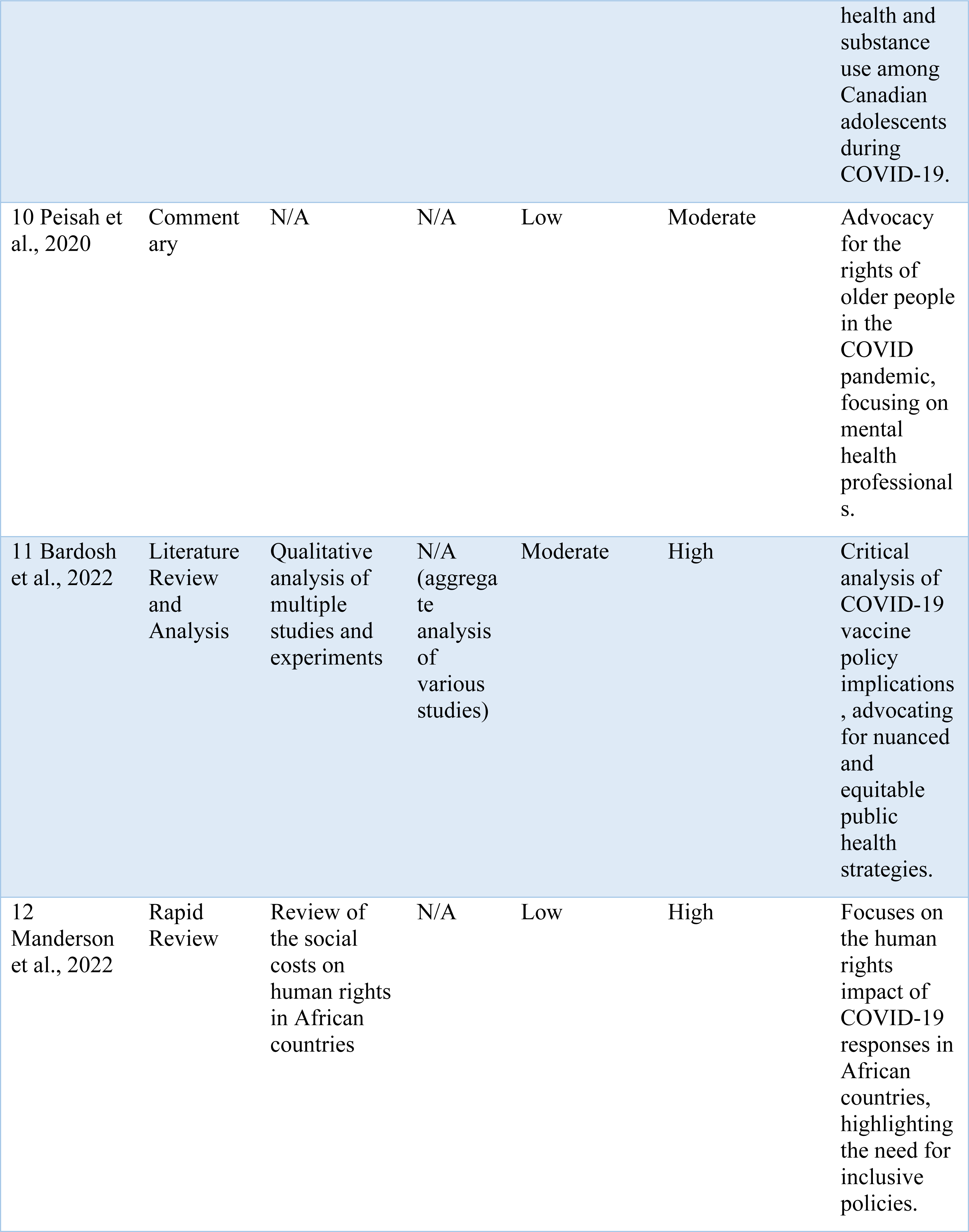
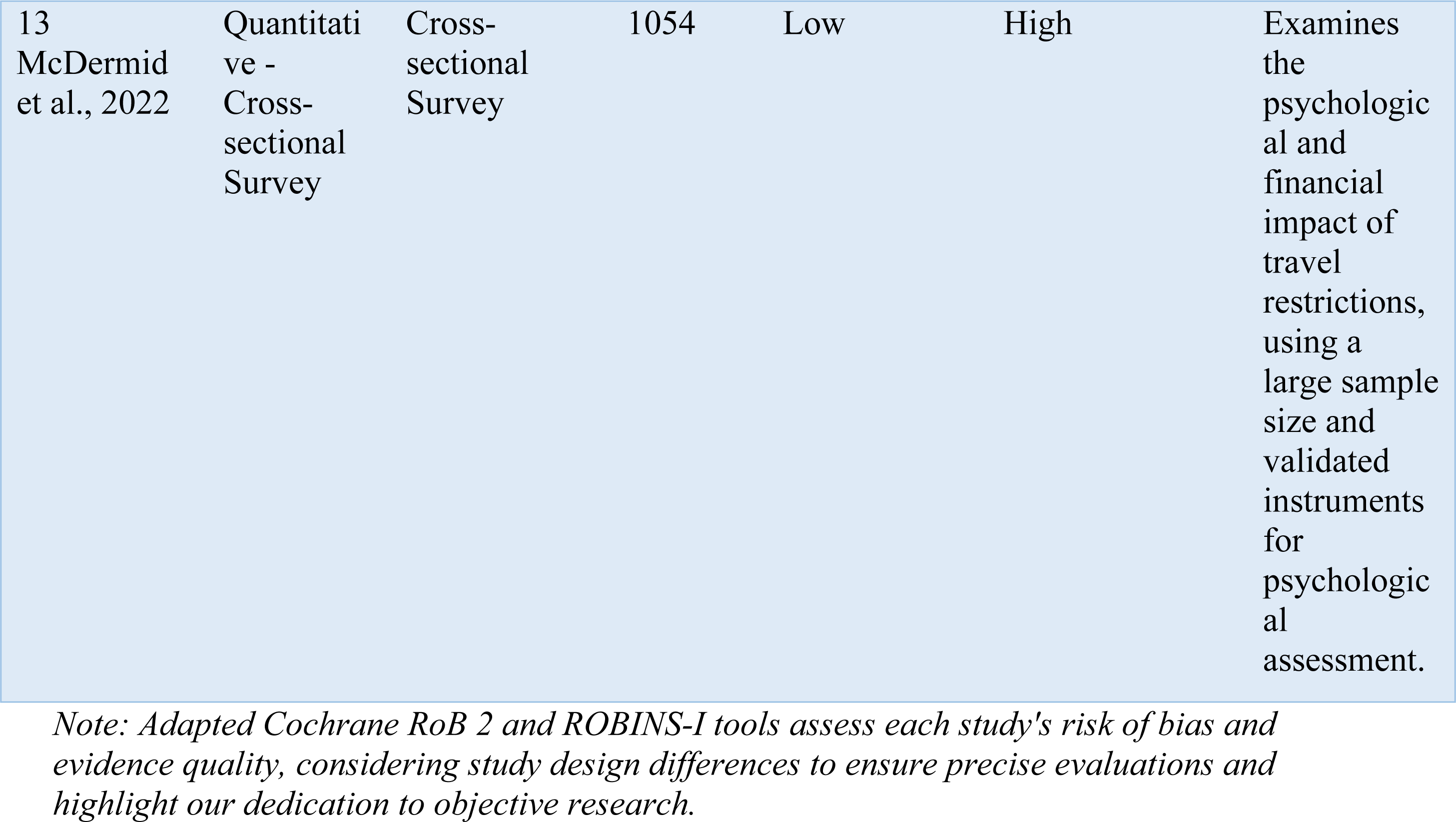
Risk of Bias & Quality Assessment of Studies.

| Study | Study Type | Study Design | Sample Size | Risk of Bias | Quality of Evidence | Notes on RoB and Quality Assessment |
| --- | --- | --- | --- | --- | --- | --- |
| 1 D'Cruz & Banerjee, 2020 | Review Article | Advocacy Review | N/A (Review) | Moderate | Moderate/High | Systematic advocacy review, grounded in research and frameworks, highlighting older adults' marginalization during COVID-19. Offers valuable human |

|  |  |  |  |  |  | rights insights. |
| --- | --- | --- | --- | --- | --- | --- |
| 2 Wood et al., 2023 | Quantitative - Cross-sectional Survey | Cross-sectional Survey | 374 | Low | High | Empirical data supports high evidence quality; low RoB due to robust survey methodology. |
| 3 Sanghi & Jaswal, 2022 | Review Article | Policy Review | N/A (Review) | Low | Moderate/High | Policy review with a systematic approach, offering insights into the right to mental health in India during COVID-19. |
| 4 Sekalala et al., 2020 | Review/Analysis | Review of International Policies and Actions | Global | Low | High | Comprehensive analysis of human rights in COVID-19 responses, advocating for rights-based policy-making. |
| 5 Jefferson et al., 2021 | Analytical Paper | Detailed analysis of the socio-political impacts | Focused on specific countries in the Global South | Medium | High | Medium RoB for depth of socio-political analysis, offering high evidence on |

|  |  |  |  |  |  | impacts in the Global South. |
| --- | --- | --- | --- | --- | --- | --- |
| 6 Cannon et al., 2023 | Quantitative | Structured interviews | 41 | Low | High | Structured interviews revealing high-quality evidence on intimate partner violence during COVID-19 with low RoB. |
| 7 Tampubolon, 2022 | Transdisciplinary Review and Thematic Analysis | Review of laws, regulations, international norms, and other literature | N/A | High | Moderate | Broad scope introduces a high RoB but provides moderate-quality evidence on the legal and normative aspects of COVID-19 responses. |
| 8 Suleman et al., 2020 | Commentary | N/A | N/A | Low | Low | Commentary with low RoB, offering insights into impacts on children and youth, though evidence quality is limited. |
| 9 Craig et al., 2023 | Cross-sectional study | Observational study design | 809 | Low | High | Observational design focused on mental health and substance use among Canadian adolescents during COVID-19. |
| 10 Peisah et al., 2020 | Commentary | N/A | N/A | Low | Moderate | Advocacy for the rights of older people in the COVID pandemic, focusing on mental health professionals. |
| 11 Bardosh et al., 2022 | Literature Review and Analysis | Qualitative analysis of multiple studies and experiments | N/A (aggregate analysis of various studies) | Moderate | High | Critical analysis of COVID-19 vaccine policy implications, advocating for nuanced and equitable public health strategies. |
| 12 Manderson et al., 2022 | Rapid Review | Review of the social costs on human rights in African countries | N/A | Low | High | Focuses on the human rights impact of COVID-19 responses in African countries, highlighting the need for inclusive policies. |
| 13<br>McDermid<br>et al., 2022 | Quantitative -<br>Cross-sectional<br>Survey | Cross-sectional<br>Survey | 1054 | Low | High | Examines the psychological and financial impact of travel restrictions, using a large sample size and validated instruments for psychological assessment. |

*Note: Adapted Cochrane RoB 2 and ROBINS-I tools assess each study's risk of bias and evidence quality, considering study design differences to ensure precise evaluations and highlight our dedication to objective research.*

The analytical paper had a medium risk of bias but offered high evidence due to the depth of analysis, whereas transdisciplinary reviews posed a high risk of bias because of their broad scope, which resulted in moderate evidence quality. Those studies marked as "N/A" or "TBD" were not applicable for risk of bias or quality assessment due to their unique nature. In conducting these assessments, systematic review practices and protocols were drawn upon, specifically referencing the Cochrane guidelines (24) and PRISMA guidelines (25). This was done while considering the applicability of each tool to the design of the studies being assessed. The entire procedure, including the evaluation of study quality and risk of bias, was conducted by a single researcher, as this review forms part of a master’s degree assignment. These assessments were intended to be objective and reproducible, as presented in Table 3.

### Data Analysis - Narrative Synthesis

The narrative synthesis approach was adopted to analyze and synthesize findings from the selected studies, allowing for a detailed exploration of the complex and heterogeneous nature of COVID-19’s social and psychological impacts. This method facilitated a comprehensive comparison, contrast, and combination of data across studies, drawing on thematic analysis to identify common threads, differences, and the broader implications of pandemic control measures. By integrating diverse evidence, this narrative synthesis unveils the nuanced ways in which COVID-19 control measures have influenced human behavior, mental health, and societal norms, offering critical insights into effective strategies for mitigating adverse outcomes while respecting human rights.

In this systematic review (Table 3.), a rigorous and meticulous assessment was conducted to thoroughly evaluate both the risk of bias and the overall quality of the included studies.

## Results

### Data Extraction

This section presents the key findings derived from the analysis of the included studies, shedding light on the diverse aspects of the social and psychological dynamics, human rights implications, and unintended consequences of COVID-19 control measures. Through a comprehensive narrative synthesis of the extracted data, this section provides a concise overview of the significant themes and patterns that emerged, offering valuable insights into the multifaceted impacts of the pandemic on various populations and the urgent need for rights-based approaches as presented in Table 4.

**Table 4.**
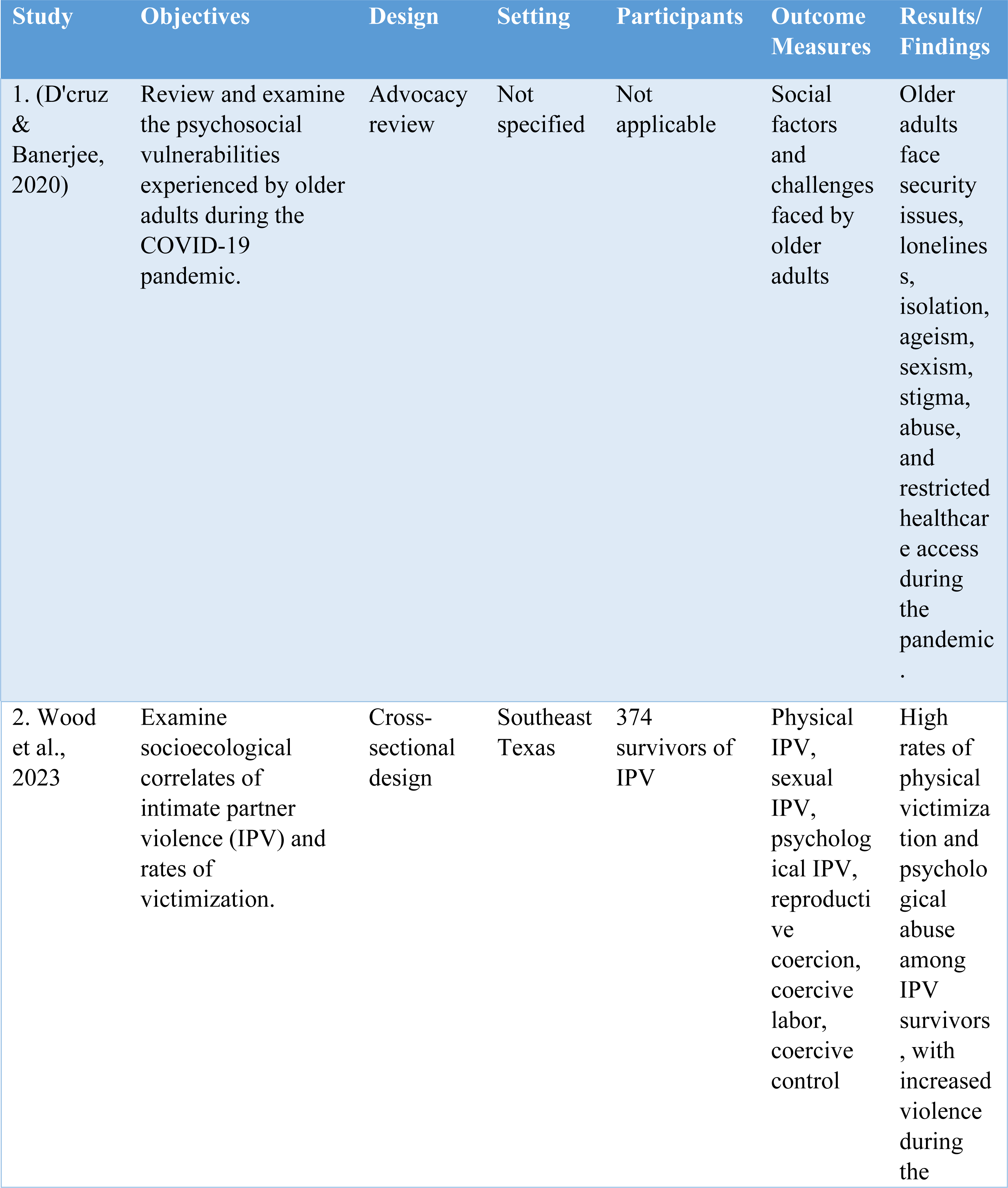
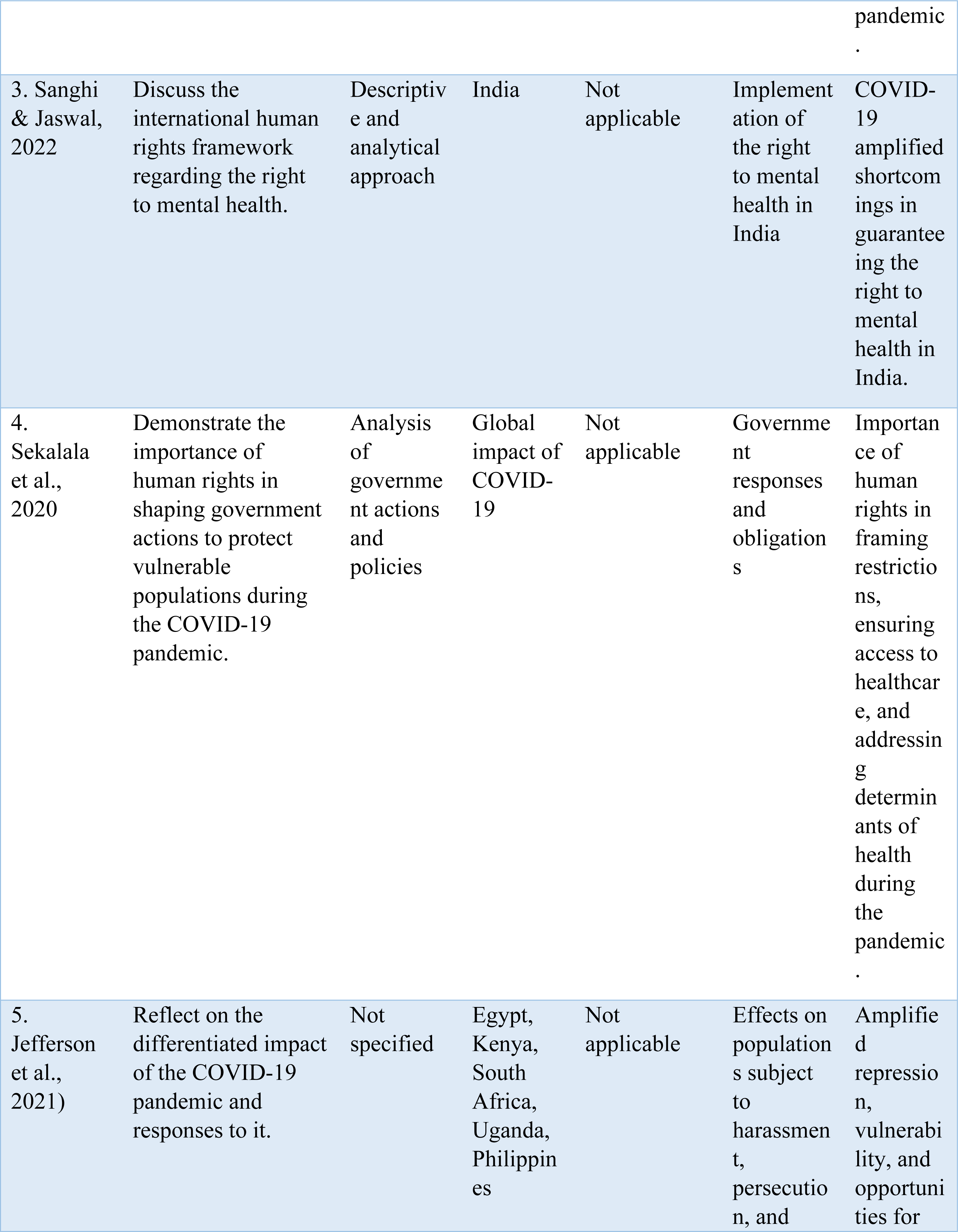
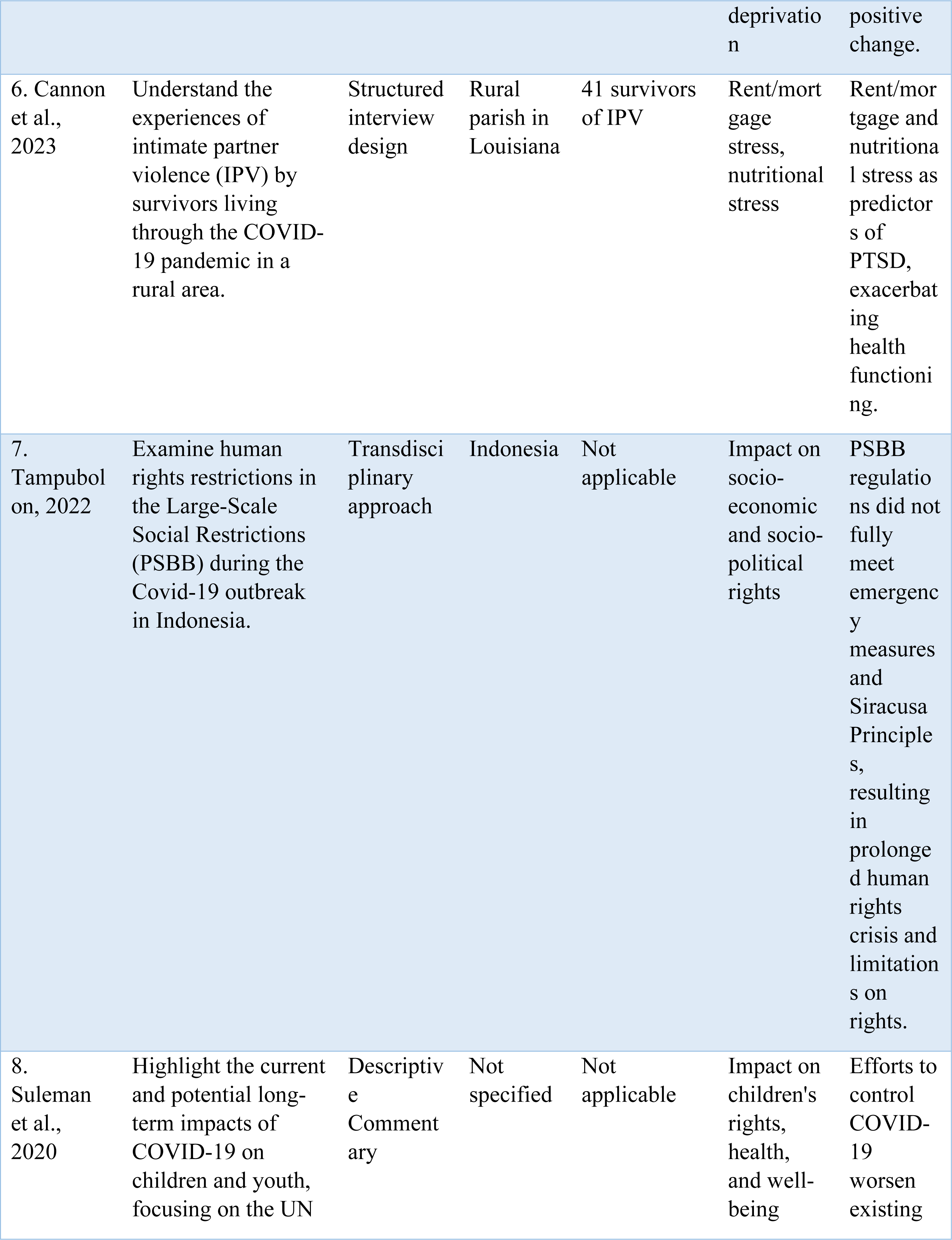
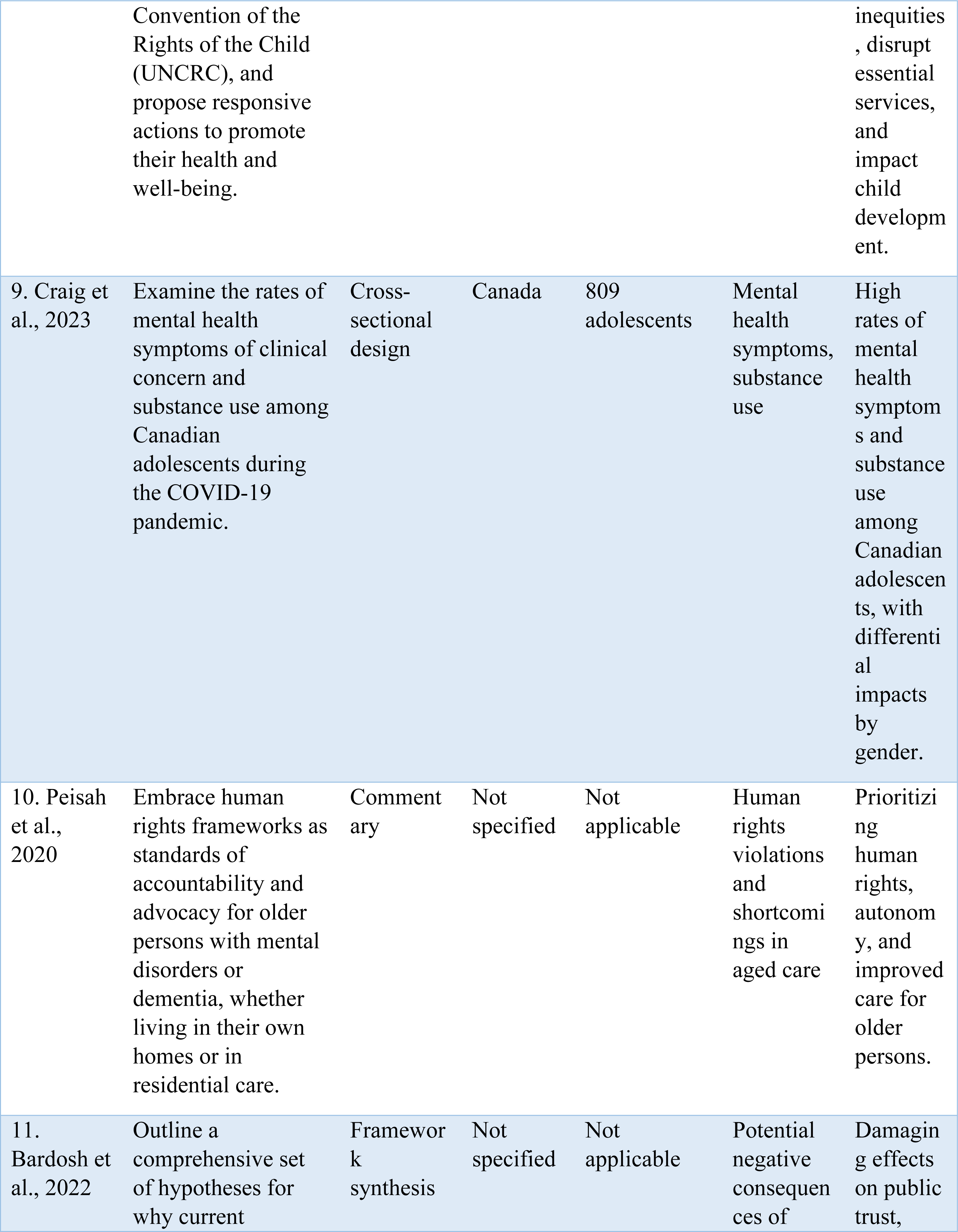
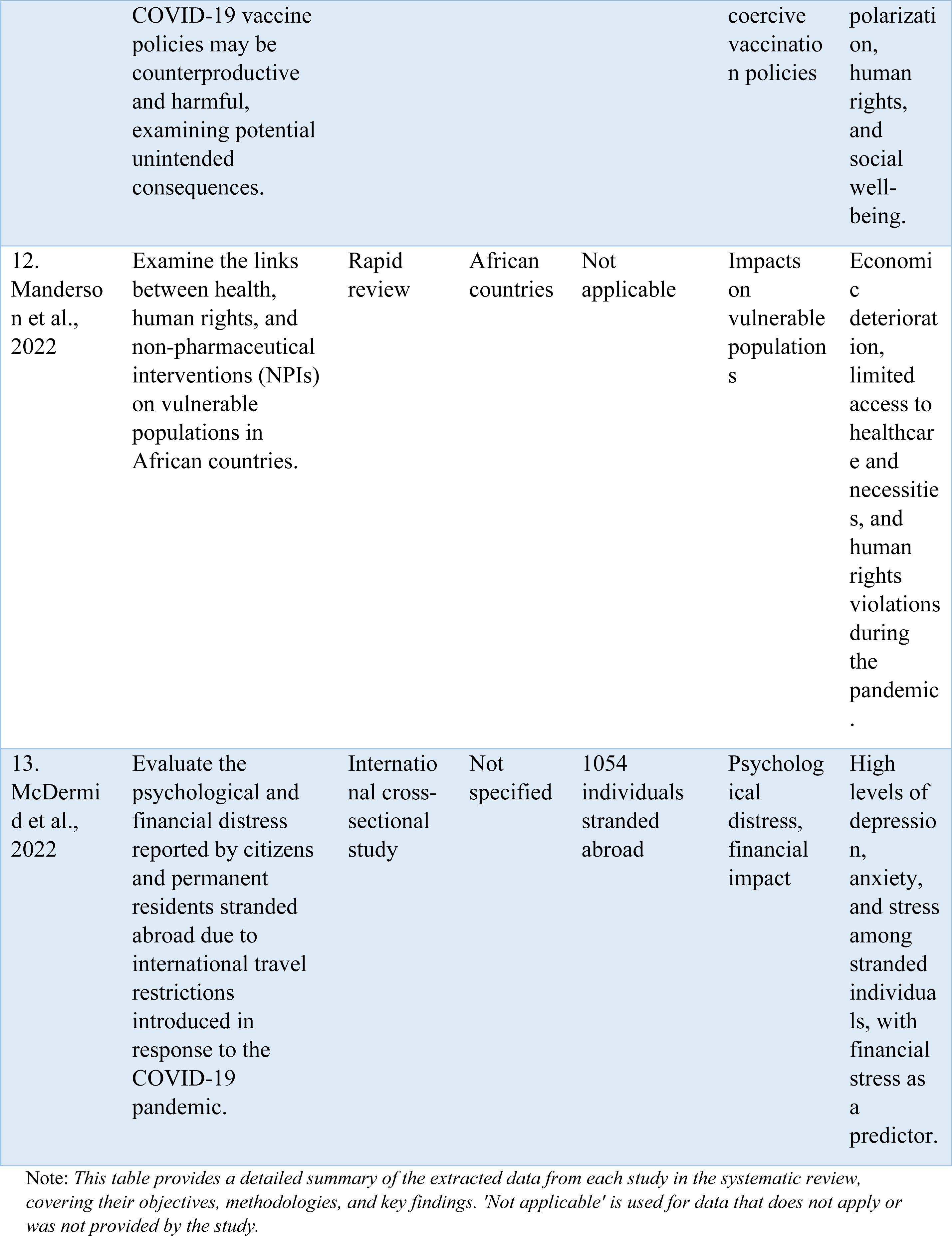
Data Extraction.

### Narrative Synthesis of Extracted Data

The COVID-19 pandemic control measures have had significant psychological and social consequences worldwide, with implications for human rights. A systematic review of 13 selected studies aimed to explore the psychological and social impact of COVID-19 control measures and their implications for human rights. The studies covered a range of topics, including the human rights and marginalization of older adults (26, 27), intimate partner violence (28), mental health rights (29), government responses (30), torture prevention (31), impacts on rural survivors of intimate partner violence (32), human rights derogation (33), the rights of children and youth (34, 35), the unintended consequences of COVID-19 vaccine policy (36), COVID-19 control measures in African countries and the psychological and financial impact of travel restriction (37, 38).

The review found that older adults were particularly vulnerable during the pandemic. They faced human rights violations and various psychosocial challenges, including security issues, loneliness, isolation, ageism, sexism, dependency, stigma, abuse, and restricted healthcare access (26, 27). Peisah et al. (27) examined the impact of the COVID-19 pandemic on the human rights of older persons, including high death rates in residential facilities, a lack of access to quality end-of-life care, restrictions on visitation rights, and resource limitations affecting care provision. D’cruz & Banerjee (26) contributed these factors to the burden faced by older adults and highlighted the marginalization and human rights deprivation experienced by this population. Recommendations were provided to mitigate marginalization and improve the implementation of the human rights framework (26).

Intimate partner violence emerged as a significant issue during the pandemic. High rates of victimization were reported, with physical and psychological abuse being the most prevalent forms. Certain demographic and economic factors, such as being older and black, were associated with higher rates of intimate partner violence (28). The study highlighted the need for a strong community response and support for survivors to address the impacts of the pandemic on intimate partner violence.

The review also focused on mental health rights during the pandemic. The study analyzed the international human rights framework and evaluated the implementation of the right to mental health in India. It found that the pandemic revealed and intensified the shortcomings in guaranteeing the right to mental health, particularly for marginalized individuals (30). Despite greater normative clarity, the practice in India and other states did not align with international human rights standards. The study emphasized the need for states to learn from the Indian experience and provided recommendations for advancing the implementation of mental health rights.

The importance of human rights in shaping government actions during the pandemic was another key theme. The review highlighted the need for government responses to be guided by human rights principles, including the necessity, proportionality, and non-discrimination of restrictions on individual rights (30). States were reminded of their obligations to ensure access to healthcare and address the underlying determinants of health. The study called for transparent policymaking, public participation, and protection of vulnerable populations in public health measures. International collaboration and assistance were also emphasized to address the global impact of the pandemic.

The review further explored the implications of COVID-19 control measures on torture prevention and human rights in the Global South (31). The crisis exacerbated existing deprivations and amplified forms of repression and vulnerability. Securitized responses to the pandemic revealed conservatism, while civil society organizations demonstrated agility and capacity for innovation. The crisis presented an opportunity for progressive change, emphasizing the relational nature of human rights and the need for anti-torture work. The study highlighted the importance of addressing human rights violations and ensuring the protection of vulnerable populations in the pandemic response.

Impacts on rural survivors of intimate partner violence were also examined in one study. The pandemic exacerbated existing concerns, with increased stress related to rent/mortgage and nutrition (32). The study found that IPV concerns may exacerbate pandemic-related concerns, further affecting the health and functioning of survivors. The findings emphasized the need for targeted support and positive coping strategies for rural IPV survivors.

The review encompassed studies examining the unintended consequences of COVID-19 vaccine policy (36), COVID-19 control measures in African countries, and the psychological and financial impact of travel restrictions (37, 38). Bardosh et al. (36) illustrated how certain vaccine policies inadvertently exacerbated inequalities and stoked vaccine hesitancy, impacting the overall effectiveness of public health efforts. This study highlights the need for a nuanced understanding of local contexts and culturally sensitive communication in vaccine distribution and policymaking. The findings of McDermid et al.’s (38) study highlighted high levels of depression, anxiety, and stress among stranded individuals. Additionally, the study identified financial stress as a significant predictor of distress in this population. The travel restrictions had severe psychological and financial repercussions for these citizens and permanent residents, underscoring the broader impacts of COVID-19 control measures on various vulnerable groups.

The review also discussed the human rights restrictions and impacts of COVID-19 control measures in Indonesia (33). The study found that the government’s response did not fully meet the emergency measurements and provisions set by international human rights frameworks. The implementation of control measures resulted in a prolonged human rights crisis, inadequate policies, manipulated data, and insufficient medical equipment. The study highlighted the consequences of ineffective regulations, including limitations on socio-economic and socio-political rights. Tampubolon’s (33) findings raised concerns about prioritizing economic considerations over saving lives and called for a balance between infection containment and human rights protection.

The rights and well-being of children and adolescents during the pandemic were explored in two studies (34, 35). Suleman et al. (34) found that efforts to control the spread of COVID-19 worsened existing inequities for marginalized children and youth. Disruptions to essential services, education, and support programs were particularly impactful for vulnerable populations. Structural inequities, xenophobia, and racism were magnified during the pandemic, further affecting the mental health and development of children and youth. Craig et al. (35) found that during the pandemic control measures, there were higher rates of mental health symptoms and substance use among Canadian adolescents, with differential impacts by gender, with over 50% of youth engaging in some form of substance use in the past 90 days and almost 20% engaging in substance use at least once a week. The studies emphasized the need for enhanced investment in services for children and the establishment of mechanisms to uphold their rights (34, 35).

The collective findings of the selected studies reveal a complex picture of the interplay between COVID-19 control measures, their psychological and social impacts, and human rights implications. The studies collectively indicate that the pandemic and the measures implemented to control it have amplified pre-existing vulnerabilities and inequalities in societies worldwide. For example, the marginalization of older adults, increased intimate partner violence, and the violation of mental health rights all point to the accentuated strain on marginalized and at-risk populations.

Contrasts also exist between studies, reflecting the complexity and multifaceted nature of the pandemic. For instance, while some studies focus on the harmful consequences of restrictive measures, others spotlight the agility and innovation of civil society organizations in response to the crisis. The common thread running through all these studies, however, is the call for a more balanced and human rights-based approach to managing the pandemic. Regardless of the specific issue examined, whether mental health rights, children’s rights, or rights related to vaccines, each study underscores the imperative for policymakers and practitioners to prioritize human rights and the well-being of individuals and groups in their responses to public health emergencies.

The systematic review provided valuable insights into the psychological and social consequences of COVID-19 control measures and their implications for human rights. It highlighted the vulnerabilities faced by older adults, survivors of intimate partner violence, and marginalized populations, as well as the importance of mental health rights and government responses guided by human rights principles. The review emphasized the need to protect the rights and well-being of children and youth and to address torture prevention and human rights violations. It called for a balance between infection containment and human rights protection, ensuring equitable access to healthcare and support services. The findings contribute to a better understanding of the multifaceted impacts of the pandemic and provide recommendations for policy and practice to mitigate the adverse effects on human rights.

The results of this systematic review offer a profound examination of the disparities and dynamics unleashed or magnified by the COVID-19 pandemic. Beyond identifying demographic disparities in the impact of COVID-19 control measures, this review illuminates the complex interplay of factors driving these disparities. It becomes apparent that the pandemic has not only deepened existing societal fractures but has also spurred community resilience and innovative responses. These findings underscore the pandemic’s role as a magnifying lens, revealing the nuances of social equity and resilience. This section calls for a nuanced reassessment of public health strategies, advocating for approaches that emphasize equity, inclusivity, and the fostering of community-led mitigation efforts.

### Thematic Analysis and Synthesis

The COVID-19 pandemic and its associated control measures have induced a complex web of psychological, social, and human rights implications, as evidenced by the comprehensive analysis of 13 selected studies. This thematic synthesis aims to unravel the interconnected themes and patterns that emerged from the diverse array of research, shedding light on the multifaceted impacts of the pandemic and the urgent need for rights-based approaches.

#### Vulnerabilities of Older Adults

Several studies underscored the heightened vulnerabilities experienced by older adults during the pandemic. They faced a multitude of challenges, including security issues, loneliness, isolation, ageism, sexism, stigma, abuse, and restricted healthcare access. The marginalized status of this demographic group was exacerbated by the pandemic, leading to profound implications for their mental health and well-being (D’cruz & Banerjee, 2020; Peisah et al., 2020).

#### Intimate Partner Violence (IPV)

The pandemic also brought to the forefront the issue of intimate partner violence, with studies reporting high rates of victimization among survivors. Physical and psychological abuse were prevalent, particularly among certain demographic and economic groups. The stressors associated with the pandemic, such as financial strain and social isolation, exacerbated the risk of IPV, highlighting the urgent need for community support and intervention strategies (Wood et al., 2023; Cannon et al., 2023).

#### Mental Health Rights

Analysis of the international human rights framework revealed significant shortcomings in guaranteeing the right to mental health, particularly in low-resource settings. The pandemic underscored the need for a rights-based approach to mental health, emphasizing the importance of equitable access to mental health services and support systems (Sanghi & Jaswal, 2022).

#### Government Responses and Human Rights

The pandemic prompted governments to implement a range of control measures to curb the spread of the virus. However, the analysis of government actions highlighted the importance of upholding human rights principles in policymaking. There was a critical need for transparent, evidence-based decision-making that prioritized the protection of human rights, particularly among vulnerable populations (Sekalala et al., 2020; Tampubolon, 2022).

#### Unintended Consequences and Human Rights Implications

Certain COVID-19 control measures inadvertently led to unintended consequences, ranging from vaccine hesitancy to economic and social disparities. Coercive vaccination policies, for instance, raised concerns about public trust, polarization, and human rights violations. Similarly, travel restrictions resulted in psychological distress and financial hardship among stranded individuals, highlighting the need for a more nuanced and rights-based approach to pandemic management (Bardosh et al., 2022; McDermid et al., 2022).

#### Impacts on Vulnerable Populations

Vulnerable populations, including children, adolescents, and rural communities, bore the brunt of the pandemic’s social and economic consequences. Disruptions to essential services, education, and support programs exacerbated existing inequities and disproportionately affected marginalized groups. The pandemic underscored the imperative of upholding the rights and well-being of these populations, emphasizing the need for targeted interventions and policy initiatives (Suleman et al., 2020; Craig et al., 2023; Manderson et al., 2022).

#### Global Implications and Human Rights Crisis

The pandemic exposed systemic weaknesses in the protection of human rights globally, with certain countries failing to meet international standards in their pandemic response. Human rights derogation, inadequate healthcare provisions, and socio-economic disparities emerged as pressing concerns, underscoring the need for international collaboration and solidarity in addressing the broader human rights crisis exacerbated by the pandemic (Jefferson et al., 2021; Tampubolon, 2022; Manderson et al., 2022).

## Discussion

This review delves into the psychological and social underpinnings of the global response to the pandemic, uncovering the layers of collective trauma experienced across communities. It examines the varied strategies employed by different societal segments to navigate pandemic-induced stressors, shedding light on the factors that bolster social cohesion and mental health resilience. This exploration reveals significant insights into post-traumatic growth and the role of social support systems in crisis times. Moreover, the analysis critically assesses the effectiveness of public health messaging and policies in bridging the trust gap among diverse populations. It suggests a forward path toward empathetic, culturally sensitive public health communication strategies that acknowledge and address the diverse needs and perspectives of global populations.

The results of this systematic review provide valuable insights into the social and psychological dynamics, human rights implications, and consequences of COVID-19 control measures. By critically analyzing and synthesizing the findings, this discussion aims to deepen our understanding of the complex issues surrounding this issue. The findings of the included studies reveal the multifaceted factors that influence individuals’ adherence to COVID-19 control measures. Drawing upon social psychology theories, we can interpret the observed patterns of behavior, considering the role of social norms, group dynamics, and psychological processes.

For instance, Wood et al. (28) emphasize the significance of social influence and perceived norms in shaping compliance with COVID-19 measures, highlighting the critical role of Social Learning Theory. This theory asserts that behaviors are learned through observation and imitation. Understanding these factors is crucial for policymakers and public health practitioners as they develop targeted interventions for future pandemics or similar events. Moreover, authorities and experts must utilize this knowledge responsibly, ensuring that interventions are implemented ethically and without exploitation.

### Critical Analysis and Synthesis

A critical examination of the human rights implications associated with COVID-19 control measures is warranted. The studies included in this review highlight potential violations of human rights, particularly among vulnerable populations such as older adults, youth, and survivors of intimate partner violence. D’cruz and Banerjee (26) shed light on the marginalization and human rights deprivation experienced by older adults during the pandemic. Similarly, Wood et al. (28) emphasize the disproportionate impact of control measures on survivors of intimate partner violence. These findings underscore the importance of upholding human rights principles, ensuring justice, and avoiding dangerous and discriminatory practices during the development and implementation of control measures.

The human rights concerns highlighted resonate with Cognitive Dissonance Theory, as individuals grapple with the tension between adhering to control measures and preserving personal freedoms. This dissonance, particularly pronounced among vulnerable groups like older adults and survivors of intimate partner violence, underscores the necessity of developing public health strategies that are not only effective but also ethically sound and rights-respecting. To further elucidate the psychological impact of COVID-19 control measures, the concept of Learned Helplessness can be applied, particularly in understanding the pervasive feelings of powerlessness and apathy among individuals facing continuous lockdowns and restrictions. This aligns with the observations made by the WHO regarding the global increase in anxiety and depression.

#### Limitations

It is important to acknowledge the limitations inherent in the included studies, which may introduce biases and limit the generalizability of the findings. Heterogeneity in study designs, sample sizes, and methodologies should be considered when interpreting the results. Additionally, reliance on self-reported data and the potential for recall bias may impact the accuracy of the findings. Future research should employ rigorous methodologies to strengthen the evidence base.

While conducting this systematic review, meticulous steps were taken to minimize potential biases and ensure a rigorous quality assessment of the included studies. Despite these efforts, it is essential to acknowledge the inherent limitations associated with the synthesis of published literature, including publication bias and the variability in study designs and methodologies. The application of the Cochrane Risk of Bias tool and ROBINS-I for non-randomized studies helped in critically evaluating each study’s reliability and validity, ensuring a balanced interpretation of the findings.

#### Implications

The implications of this study extend to various domains, including social, clinical, and research practices. From a social perspective, the findings underscore the importance of community engagement, trust-building, and transparent communication in public health interventions. Furthermore, engaging with communities can help tailor interventions to address specific cultural, social, and economic contexts. Suleman et al. (34) highlight the need for a rights-centered approach to supporting children and youth during the pandemic, emphasizing the principles outlined in the UN Convention on the Rights of the Child. This approach promotes the protection and fulfillment of children’s rights, considering their specific vulnerabilities and needs.

Clinically, healthcare providers can utilize these insights to inform person-centered care approaches. The mental health implications of the pandemic have been significant, with individuals experiencing heightened levels of distress, anxiety, and depression. Sanghi and Jaswal (29) discuss the impact of the pandemic on mental health rights, calling for the implementation of the right to mental health in India. Addressing the mental health needs of vulnerable populations, including older adults and survivors of intimate partner violence, requires a comprehensive and integrated approach that prioritizes access to quality mental health services, support systems, and advocacy for their rights.

From a research standpoint, interdisciplinary collaborations, rigorous methodologies, and long-term monitoring of the social and psychological impacts of pandemics are essential. Bardosh et al. (36) highlight the importance of incorporating a One Health approach in studying the consequences of COVID-19 control measures. This approach recognizes the interconnectedness of human, animal, and environmental health and considers the social determinants of health in designing interventions. Longitudinal studies are needed to investigate the long-term consequences of the pandemic control measures on mental health, well-being, and human rights. Understanding the long-lasting impacts of the pandemic is crucial for developing evidence-based strategies to mitigate the adverse effects and support individuals and communities in their recovery.

### Future Directions

Moving forward, further exploration is warranted in several areas identified by this systematic review. Researchers should delve deeper into understanding the nuanced mechanisms that influence compliance and coercive behaviors and their misuse, considering the interplay between social, psychological, and cultural factors. Longitudinal studies can provide transparent insights into the temporal dynamics of adherence and the factors that contribute to positive and sustained behavior change. Additionally, investigations into the effectiveness of interventions aimed at promoting compliance and protecting human rights can inform future policy and practice. It is essential to adopt an evidence-based approach to decision-making and continually evaluate the impact and ethical implications of control measures.

This systematic review, informed by the insights from the selected studies, sheds light on the complex issues surrounding the social and psychological dynamics, human rights implications, and consequences of COVID-19 control measures. It emphasizes the importance of understanding factors influencing adherence, upholding human rights, and addressing the limitations inherent in the included studies. The effects of this study extend to various domains, emphasizing the need for community engagement, transparency, person-centered care, interdisciplinary collaborations, and further research. By addressing these issues, we can strive for more effective and reasonable approaches to managing the pandemic while safeguarding the human rights and well-being of individuals.

During this systematic review, rigorous steps were taken to minimize biases and ensure thorough quality assessment. Despite these efforts, limitations persist due to publication bias and variability in study designs. Utilizing tools like the Cochrane RoB tool and ROBINS-I helped assess study reliability, but addressing biases in future research and examining diverse socio-demographic contexts is essential for a nuanced understanding of COVID-19 control measure impacts. This acknowledgment emphasizes the need for cautious interpretation and suggests directions for future inquiry.

### Policy recommendations

The COVID-19 pandemic has unveiled significant socio-economic and human rights challenges, necessitating a comprehensive and multifaceted policy response. Based on the findings of our systematic review, we propose a series of policy recommendations aimed at mitigating these challenges. These recommendations are designed to guide policymakers in formulating strategies that are not only effective in controlling the spread of COVID-19 but also equitable and respectful of human rights. Below, we outline these policy recommendations, providing a rationale for each, identifying the target populations, and suggesting practical implementation strategies.

#### Ensuring Equitable Access to Healthcare and Support Services

Rationale: Vulnerable and marginalized communities have disproportionately borne the brunt of the pandemic’s impact, highlighting the urgent need for equitable access to healthcare and support services.

Target Population: This policy focuses on vulnerable populations, including those with limited access to healthcare, the elderly, low-income families, and marginalized communities.

Implementation Strategies:

- Develop outreach programs specifically designed to reach underserved communities, ensuring they are informed about and can access necessary healthcare services.
- Implement subsidies or financial assistance programs to reduce the financial barriers to healthcare access, making testing, treatment, and vaccination more accessible to all.

#### Enhancing Transparency and Public Participation

Rationale: Trust and compliance with public health measures can be significantly improved through transparent decision-making and active engagement of the public and stakeholders in the policy process.

Target Population: The general public, including stakeholders such as healthcare providers, community leaders, and patient advocacy groups.

Implementation Strategies:

- Establish a schedule of regular public briefings to explain decision-making processes, current measures, and future plans.
- Create and promote platforms for public feedback and contributions to policy decisions, ensuring a broad range of voices are heard and considered.

#### Fostering Psychological Resilience and Social Support

Rationale: The pandemic has led to widespread psychological distress and social isolation. Addressing mental health challenges is essential for societal well-being.

Target Population: Individuals experiencing mental health challenges, including those facing increased anxiety, depression, or isolation due to the pandemic.

Implementation Strategies:

- Expand access to mental health counseling and support groups, both in-person and through telehealth services.
- Launch community-based mental health initiatives to promote resilience, wellness, and social connection, tailoring interventions to meet local needs and cultural contexts.

#### Upholding Human Rights in Enforcement of Control Measures

Rationale: Protection of human rights is paramount, particularly in ensuring that enforcement of control measures does not lead to excessive use of authority, discrimination, or other abuses.

Target Population: All individuals affected by COVID-19 control measures, with particular attention to safeguarding the rights of the most vulnerable.

Implementation Strategies:

- Provide training for law enforcement personnel and others involved in enforcement activities on human rights principles and ethical enforcement practices.
- Establish oversight mechanisms and channels for reporting and addressing grievances related to the enforcement of control measures, ensuring accountability and redress.

These policy recommendations, grounded in the evidence from our systematic review, offer a roadmap for navigating the complex interplay between public health imperatives and the need to safeguard socio-economic well-being and human rights during the COVID-19 pandemic.

Implementing these recommendations requires a concerted effort from governments, civil society, and international organizations. Adjustments and customizations may be necessary to align with local contexts, resource availability, and specific challenges. However, by adhering to these principles, policymakers can enhance the effectiveness, equity, and acceptability of their responses to the pandemic, ultimately contributing to a more resilient and just society.

## Conclusion

In conclusion, this systematic review articulates a visionary perspective for the evolution of public health frameworks in the aftermath of the COVID-19 pandemic. Advocating for an integrated approach to health that harmonizes physical well-being, psychological health, and social justice, it calls for the establishment of a robust, adaptive public health infrastructure. This infrastructure should not only be capable of effectively responding to immediate crises but also proactive in nurturing a resilient society equipped to face a broad spectrum of future challenges. The review highlights a pressing need for further research to explore the mechanisms of social resilience and the integration of human rights within health crisis management. By emphasizing these areas, the manuscript aims to contribute to a transformative dialogue on reimagining public health paradigms for a more equitable and resilient future.

This discussion provides a comprehensive analysis and interpretation of the study’s findings, grounded in research evidence and theoretical frameworks. It critically examines the complex dynamics of COVID-19 control measures, their implications for human rights, and the social and psychological factors influencing compliance behaviors. The synthesis of the selected studies sheds light on the multifaceted nature of the pandemic and its far-reaching consequences for individuals and communities. The review’s key findings suggest that adherence to COVID-19 control measures is significantly influenced by various social and psychological factors and that these measures can have profound human rights implications, particularly for vulnerable populations.

The limitations inherent in the included studies, such as variations in methodologies and potential biases, should be taken into consideration when interpreting the findings. Nonetheless, the collective evidence highlights the need for a balanced approach that upholds human rights while addressing public health objectives. It emphasizes the importance of community engagement, trust-building, and transparent communication in designing and implementing control measures.

The conclusions of this discussion extend to multiple domains. From a social perspective, it underscores the significance of fostering community resilience, ensuring equitable access to resources, and addressing social determinants of health. Clinically, healthcare providers should be attuned to the unique needs and vulnerabilities of different populations, incorporating a human rights-based approach into their practice. In terms of research, there is a call for further interdisciplinary collaborations, rigorous methodologies, and long-term monitoring to deepen our understanding of the social, psychological, and human rights dimensions of pandemics.

Looking ahead, integrating human rights principles into pandemic response strategies is crucial for creating more inclusive, equitable, and effective interventions. Policymakers, researchers, and practitioners must work together to navigate the complex challenges posed by the pandemic while safeguarding the rights and well-being of individuals. By drawing on the insights from this discussion, we can forge a path towards a more resilient and rights-based approach to public health crises.

## Data Availability

This systematic review analyzes data derived entirely from publicly available sources. The findings and conclusions presented are based on a comprehensive review of peer-reviewed articles, reports, and official documents, which have been previously published and are accessible in the public domain. The review methodology, including detailed search strategies, selection criteria, and theoretical frameworks, is thoroughly outlined within the manuscript, specifically in the Methods section. Specific data extraction tables, risk of bias assessments, and narrative syntheses are provided in the Results and Discussion sections, offering full transparency on the data synthesized in this review. The databases searched during the study include ProQuest, Science Direct, PsycINFO/PsycNET, and PubMed. Due to the nature of this research as a systematic review, no primary dataset was generated. Hence, the original articles and sources can be accessed directly through the respective databases and journals in which they are published. For any queries regarding the search methodology or to request the search strategies employed (presented in the manuscript), interested researchers can contact the corresponding author. No proprietary or confidential data were used in this study. As such, there are no legal or ethical restrictions on the data, and no exemptions to the data availability policy are requested.

## Acknowledgments

I extend my sincere appreciation to Liverpool John Moores University for providing the academic environment and resources that facilitated the development of this systematic review. While this project was independently conducted, the foundational knowledge and research skills acquired during my time at the university were invaluable. I am also grateful for the various open access resources and databases that made this research possible.

## Disclaimer

The views and interpretations in this manuscript are those of the author and do not necessarily represent those of Liverpool John Moores University.

## Funding

This research did not receive any specific grant from funding agencies in the public, commercial, or not-for-profit sectors.

## Ethics Approval

This systematic review was conducted based on secondary data analysis, involving no direct human subjects, human material, or human data collection. The research was conducted in accordance with the ethical standards of academic research and the Declaration of Helsinki.

## Data Availability Statement

The data that support the findings of this study are derived from publicly available sources. Detailed search strategies, selection criteria, and theoretical frameworks are extensively outlined within the Methodology section of the manuscript. Specific data extraction tables, risk of bias assessments, and narrative syntheses can be found in the Results and Discussion sections. The original .ris files used for the systematic literature search, representing a foundational aspect of the research process.

The data underlying this study come from detailed database searches, including Science Direct, PubMed, PsycNET, and ProQuest. Due to institutional access limitations, ProQuest platform search history and sharable links cannot be accessed, although ris file exists. However, I maintain full access to search histories and methodologies for Science Direct, PubMed, and PsycNET. This manuscript includes search strategies, keywords, filters, and outcomes for each database, demonstrating my commitment to transparency and reproducibility. These details can be provided upon request.

## Notes

### Competing Interest Statement

The authors have declared no competing interest.

### Author Declarations

This manuscript, a systematic review, synthesizes existing research findings and does not involve new human participant data. All studies included were assessed for adherence to ethical standards as reported in their publications. The review relied on publicly available data from studies with ethical approval by respective institutional review boards or ethics committees.

